# The COVID-19 Critical Care Consortium observational study: Design and rationale of a prospective, international, multicenter, observational study

**DOI:** 10.1101/2020.05.29.20115253

**Authors:** Gianluigi Li Bassi, Jacky Y. Suen, Adrian G. Barnett, Amanda Corley, Jonathan E. Millar, Jonathon P. Fanning, India Lye, Sebastiano Colombo, Karin Wildi, Samantha Livingstone, Gabriella Abbate, Samuel Hinton, Benoit Liquet, Sally Shrapnel, Heidi J. Dalton, John F. Fraser

## Abstract

**Importance:** There is a paucity of data that can be used to guide the management of critically ill patients with coronavirus disease 2019 (COVID-19). Global collaboration offers the best chance of obtaining these data, at scale and in time. In the absence of effective therapies, insights derived from real-time observational data will be a crucial means of improving outcomes.

**Objective:** In response to the severe acute respiratory syndrome coronavirus 2 (SARS-CoV-2) pandemic, a research and data-sharing collaborative has been assembled to harness the cumulative experience of intensive care units (ICUs) worldwide. The resulting observational study provides a platform to rapidly disseminate detailed data and insights.

**Design:** The COVID-19 Critical Care Consortium observational study is an international, multicenter, prospective, observational study of patients with confirmed or suspected SARSCoV-2 infection admitted to ICUs.

**Setting:** This is an evolving, open-ended study that commenced on January 1^st^, 2020 and currently includes more than 350 sites in over 48 countries. The study enrolls patients at the time of ICU admission and follows them to the time of death, hospital discharge, or 28 days post-ICU admission, whichever occurs last.

**Participants:** All subjects, without age limit, requiring admission to an ICU for SARS-CoV-2 infection, confirmed by real-time polymerase chain reaction (PCR) and/or next-generation sequencing or with high clinical suspicion of the infection. Patients admitted to an ICU for any other reason are excluded.

**Main outcomes and measures:** Key data, collected via an electronic case report form devised in collaboration with the ISARIC/SPRINT-SARI networks, include: patient demographic data and risk factors, clinical features, severity of illness and respiratory failure, need for non-invasive and/or mechanical ventilation and/or extracorporeal membrane COVID–19 CCC observational study protocol oxygenation (ECMO), and associated complications, as well as data on adjunctive therapies. Final outcomes of in-hospital death, discharge or continuing admissions at 28 days.

**Discussion:** This large-scale, observational study of COVID-19 in the critically ill will provide rapid international characterization. Open-ended accrual will increase the power to answer hypothesis-led questions over time. Several sub-studies have already been initiated, examining hemostasis, neurological, cardiac, and long-term outcomes.

## INTRODUCTION

The world is currently witnessing a viral pandemic. Cases of atypical pneumonia first emerged in Wuhan, China, in December 2019 ^1^. Investigation has identified the cause as a novel betacoronavirus, ultimately named severe acute respiratory syndrome coronavirus-2 (SARS-CoV-2)^2^. The virus, and the disease it causes – COVID-19 – has since spread internationally. The World Health Organization declared the outbreak a “Public Health Emergency of International Concern” on the 30^th^ of January, 2020, and a “pandemic” on the 12^th^ of March. There have now been more than 5.4 million confirmed infections globally, resulting in 340,000 deaths (as of the 26^th^ of May, 2020)^3^.

### SARS-CoV-2, COVID-19, and critical illness

The mortality rate of COVID-19 among patients admitted to the intensive care unit (ICU) has been reported to be as high as 60%^4^–^7^. Early data and clinical experience indicate that this is caused primarily by acute hypoxemic respiratory failure (AHRF)^8^–^10^. These same data have also prompted some authors to suggest that the pathobiology of COVID-19 – associated AHRF may differ from that of Acute Respiratory Distress Syndrome (ARDS)^11^,^12^. This assertion hinges on reports of patients with severe COVID-19 associated AHRF and high pulmonary compliance, a presentation not thought to be typical of ARDS. Much has also been made of the high incidence of thromboembolic events in critically ill patients ^13^, ^4^. However, many reports are limited by either small numbers of patients or by geographic restrictions. These fail to account for variations in practices or for the variations between countries in patient, systemic, and organizational factors. Consequently, much of our current practice is driven by anecdotal cases or by limited case series.

### Rationale for developing a worldwide registry of COVID-19 patients admitted to ICUs

We aim to improve conclusions robustness regarding the management, interventions and treatment of critically-ill COVID-19 patients around the world. We aim to do this by utilizing combined data sets which detail a wide variety of patients entering the ICU at multiple stages of COVID-19 illness from diverse geographic locations. This ongoing research effort will aid in developing best practices based on evidence from a wide variety of ICUs throughout the world. This is especially important as there is currently a paucity of evidence-based guidelines and limited clinical resources globally. This data will also aid decision-making of clinicians working in healthcare systems that are currently managing or yet to face a surge in COVID-19 cases.

## MATERIALS AND METHODS

### Study design

This is an international, multicenter, prospective, observational study. The study protocol v. 1.2.8 appears in [Supplement 1].

The inclusion criteria are: (1) clinically suspected or laboratory-confirmed SARS-CoV-2 infection (by real time PCR and/or next generation sequencing), and (2) admission to an ICU. Patients admitted to an ICU for a reason other than SARS-CoV-2 infection are excluded. Patients of all ages from infants through adults can be enrolled into the study.

### Enrolment and participating sites

This study commenced on January 1^st^, 2020. There is no fixed end date for the study. Currently, 350 centers are included, spanning 48 countries [Supplement 2], coordinated by regional leads and assistants [Supplement 3] and the operating team at the coordinating site [Supplement 3]. Co-enrolment with other studies,including interventional trials, is permitted.

### Outcome measures

A summary of variables recorded by the study case report form (CRF) is presented in Table 1.

**Table 1.**
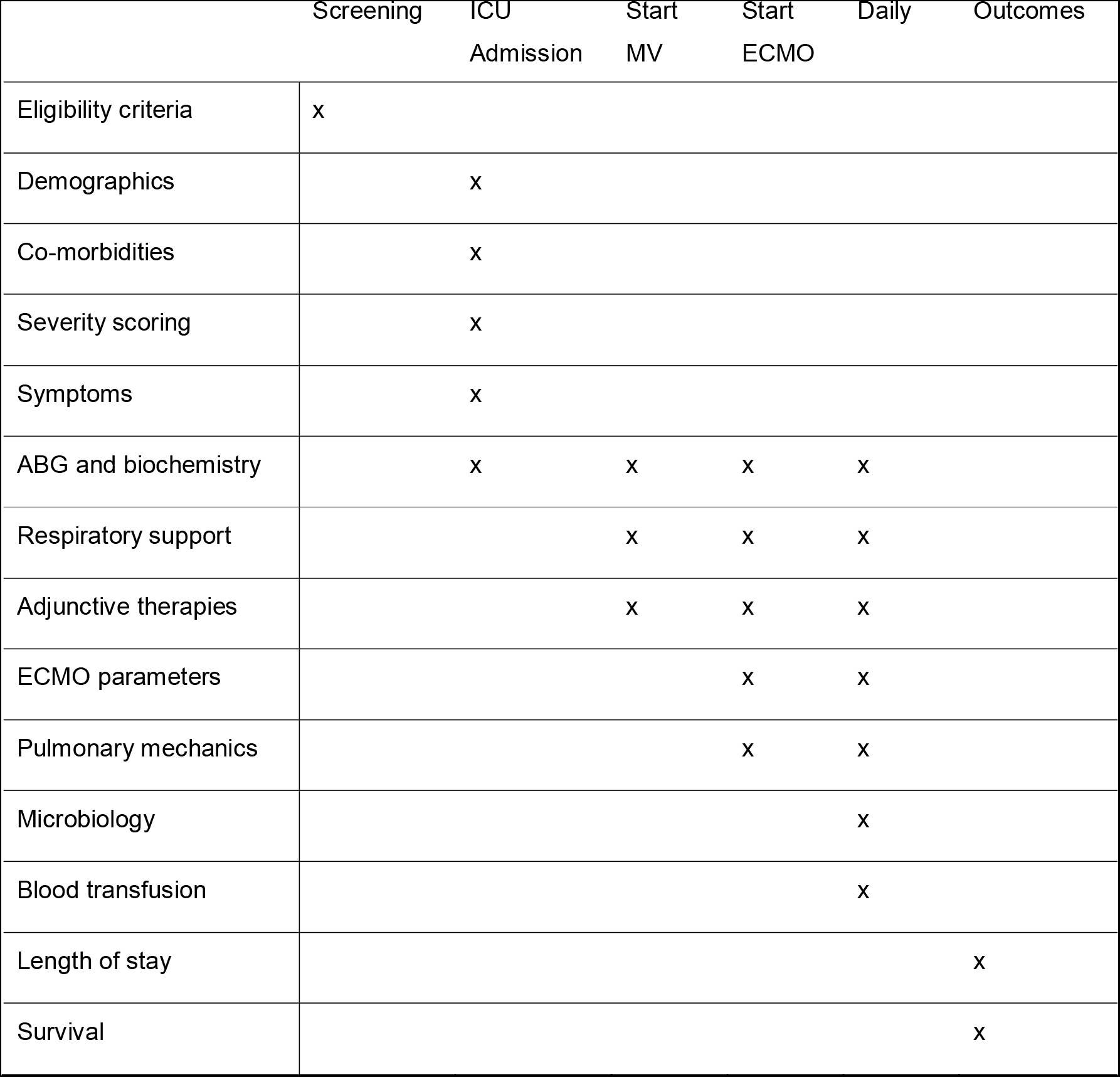
Assessment schedule. MV – mechanical ventilation; ECMO – extracorporeal membrane oxygenation; ABG –xg arterial blood gas.

|  | Screening | ICU<br>Admission | Start<br>MV | Start<br>ECMO | Daily | Outcomes |
| --- | --- | --- | --- | --- | --- | --- |
| Eligibility criteria | x |  |  |  |  |  |
| Demographics |  | x |  |  |  |  |
| Co-morbidities |  | x |  |  |  |  |
| Severity scoring |  | x |  |  |  |  |
| Symptoms |  | x |  |  |  |  |
| ABG and biochemistry |  | x | x | x | x |  |
| Respiratory support |  |  | x | x | x |  |
| Adjunctive therapies |  |  | x | x | x |  |
| ECMO parameters |  |  |  | x | x |  |
| Pulmonary mechanics |  |  |  | x | x |  |
| Microbiology |  |  |  |  | x |  |
| Blood transfusion |  |  |  |  | x |  |
| Length of stay |  |  |  |  |  | x |
| Survival |  |  |  |  |  | x |

#### Data collection methods

Streamlined data-collection instruments and procedures are used to minimize the workload at study centers. Data collection begins at the time of hospital admission using the International Severe Acute Respiratory and Emerging Infection Consortium (ISARIC) and Short Period Incidence Study of Severe Acute Respiratory Illness (SPRINT-SARI) data tools (https://isaric.tghn.org/COVID-19-CRF/). Data collection for the COVID-19 CCC observational study commences at the time of a patient’s admission to an ICU, using a study specific adaptation of the ISARIC/SPRINT-SARI COVID-19 CRF [Supplement 4]. Figure 1 outlines the schedule of assessments used for patients included in the COVID-19 CCC study. De-identified study data are collected and managed using the REDCap electronic data capture tool hosted at the University of Oxford, United Kingdom^15^. Of note, an optional, interactive augmented data collection has been implemented through a platform developed specifically for the study by Amazon Web Services Australia (AWS, Sydney, Australia). A physical device and associated software tools assist with de-identified data collection and their transfer to the REDCap database. This approach has no impact on the ownership of data, which remains with the individual site. Full encryption is used, beginning from data ingestion into the Amazon cloud, through to transfer to the REDCap web application. Data will not be used for any purpose other than those described in the study protocol. Each site’s principal investigator is responsible for ensuring data integrity. Regular written and web-based training is provided. In countries unable to upload data into a centralized database, the ability to retain a local database on a national server is available, with aggregated anonymized data exported centrally for analysis.

**Figure 1.**
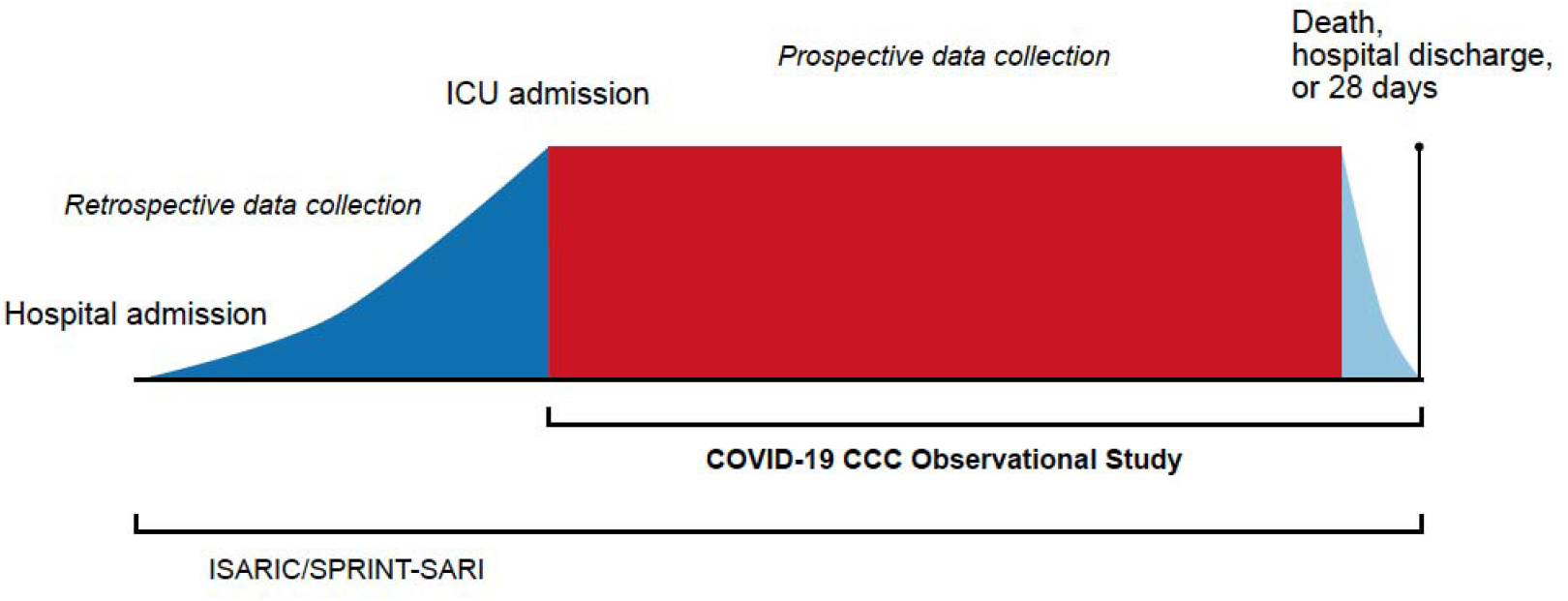
Schematic study overview. The study ends at death, hospital discharge/transfer, or 28 days, whichever occurs latest

#### Inter-hospital transfer

If a patient is transferred from a facility participating in the COVID-19 CCC and ISARIC/SPRINT-SARI to another participating center, the patient’s previously allocated unique identifier transfers with them. However, sites will not have access to study data collected outside their hospital. It is the responsibility of each hospital to enter data pertaining to their component of the patient’s hospital admission. If a patient is transferred to a non-participating hospital, there will be no further data collection. All sites will be asked to include a COVID-19 CCC and ISARIC/SPRINT-SARI study information sheet in any outgoing patient’s documentation.

#### Data management

Several procedures are in place to optimize data quality and completeness. These include: (1) a detailed data dictionary, (2) quality assurance within the data management system, (3) quality assurance of key variables within the CRF, and (4) regular written and web-based training for local study investigators. A compendious CRF is fundamental to the success of this study. Extensive efforts have been made to limit data collection to essential variables. It is hoped that this will contribute to more complete data entry with a reduced burden on participating centers. Information that is not available to the investigator will not be treated as missing, and no assumptions will be made for missing data. An audit will be conducted on a randomly selected sample (approximately 5%) of cases. In-person site visits will not be feasible, given the nature of the study and pandemic. Sub-study projects will be accessed via the main CRF platform. Specific extensions will be used to collect additional variables, limiting the overall burden on data collectors, but allowing centers involved in sub-studies to enter data in the single REDCap format.

#### Data access

The coordinating team will have access to all collected data to assure integrity, provide oversight, and conduct the main study analyses. Individual sites will have access to all the data they collect. A multinational steering committee [Supplement 1] oversees registry operations worldwide and approves investigator-initiated or site-specific sub-studies, external requests for data, and reviews suggestions by participants. To date, several sub-studies have been initiated focusing on the impact of COVID-19 on the brain, heart, kidneys, management and risks of ECMO, coagulation and thrombosis risks and long-term effects, all involving multi-center participation. Once approval is obtained, relevant de-identified data will be made available. It is anticipated that, following study completion, all de-identified data will be made open access.

#### Dissemination

Due to the evolving nature of the pandemic and the uncertainty surrounding its impact, this study was designed to be responsive to the international call for swift characterization of COVID-19 patients. Hence, in collaboration with University of Queensland and extramural collaboration with IBM Australia (St. Leonard’s,Australia), a web-based dashboard has been developed to provide relevant data and descriptive statistics to international collaborators in real-time.

## Ethical considerations

Chief investigators and the study management team are responsible for ensuring that the study is conducted in accordance with both the protocol, Declaration of Helsinki and the Principles of Good Clinical Practice. The study management team will continue to work with local principal investigators to ensure that the study adheres to all relevant national regulations, and that the necessary approvals are in place before a site may contribute data. The principal investigator at each site is responsible for maintaining a securely-held enrolment log, linking each patient’s hospital record number with the COVID-19 CCC study number, if required. The original protocol and subsequent amendments will be translated into the main language of the collaborating institutions and submitted for institutional review board approval or an equivalent. Patients will not be enrolled under the conditions of an amended protocol, until after approval has been granted.

It is expected that this study will not require informed consent in most jurisdictions. This study is, in effect, a large-scale clinical audit, as all data are collected routinely. This may justify a waiver of consent. Any jurisdiction that deems informed consent necessary may use forms provided on our website (https://www.elso.org/COVID19/ECMOCARD.aspx). Within such jurisdictions, patients who meet the eligibility criteria will be approached directly. If this is not possible, due to the patient’s incapacity, a model of retrospective or representative consent may be used, per local requirements.

### Statistical considerations

Initial characterization will be descriptive, including all eligible patients at participating centers enrolled within defined timeframes. Where analysis is hypothesis-driven, sample size calculations and power analysis (where appropriate) will depend on the specific outcome or endpoint under consideration and will be pre-defined. Results that aim to show an association or test a hypothesis will include 95% confidence intervals. These intervals and associated means will be interpreted in terms of their clinical and statistical significance, and discussion may include whether a comparison is under-powered.

For discharge, mortality, and length-of-stay outcomes, we will use a survival analysis with competing risks approach ^16^. We will graphically depict the risks of death and discharge over time using cumulative incidence plots. We will estimate which patient variables influence the risk of death and discharge using Cox regression, with separate models for death and discharge. In addition to Cox models, we will construct non-linear predictive models for both outcomes using Random Forest models, which will be externally validated on a hold-out test set. Comparison of the predictive performance of both the Cox regression and Random Forest modelling approaches will be made using: (1) a Brier score ^17^, (2) area under the receiver operating characteristic (ROC) curves using a 2-sided DeLong test, and (3) calibration plots, characterized by visual inspection and reporting of slope and intercept^17^. For the Random Forest models, a Shapley Tree Explainer will be used to identify variables that are highly predictive of each outcome^18^. This analysis will follow the Transparent Reporting of a Multivariable Prediction Model for Individual Prognosis or Diagnosis (TRIPOD) reporting guideline for prediction model development and validation^19^.

To show within-patient trends, we will plot continuous longitudinal variables over time using line plots. We will summarize each trend using daily averages and will estimate trends over time and the influence of patient variables using a linear mixed model with a random intercept per patient to control for repeated data. For binary variables, we will use panel bar charts to show the average change over time, and will model these variables using a generalized linear mixed model with a binomial distribution. A smooth estimation using cubic spline will be explored to estimate potential non-linear trends of the continuous longitudinal variables and binary variables.

## DISCUSSION

Herein we have described the rationale and design of an international, multicenter, observational registry of COVID-19 patients admitted to an ICU. To date, the characterization of patients admitted to ICUs with COVID-19 has been limited to national or single-center series. This study, using a large collaborative network, attempts to overcome the limitations induced by small patient numbers and geographic restrictions, by providing real-time global data. In a pandemic of an emerging pathogen, high-quality, real-time information is crucial to guide an optimal response. The speed of this response and cumulative experience of ICUs worldwide offer the best framework for determining evidence-based best practices and, therefore, improving outcomes for those requiring critical care.

The design of the COVID-19 CCC study has several strengths. First, the care of patients admitted to the ICU, specifically those who are mechanically ventilated, is dependent on regional resources and may vary ^20^,^21^. This potential heterogeneity is mitigated by the international composition of the consortium. Second, the study leverages novel data acquisition methods, which may improve and expedite data collection. Third, the registry-based, collaborative, and open-source approach of the study lends itself to the conduct of multiple prospective sub-studies. Fourth, the study incorporates the provision of a web-based dashboard, which provides real-time data in an accessible format.

### Limitations

Patients will not receive identical treatments and care. While this will limit some aspects of data analysis, it will also give breadth to the scope of the investigation, as data on laboratory and patient characteristics, interventions and adjunct therapies, and outcomes will be available.

This study relies on clinicians and support staff to accurately record data during a time of increased patient influx and ICU workload, raising concerns over data input error and completeness. To overcome this, coordinators at each site have access to regular training, as well as ‘drop-in’ query sessions on-line.

## Conclusions

This study will provide inclusive global characterization of critically ill patients with COVID-19. As the study is open-ended, continued data accrual will result in increased power to answer hypothesis-led questions over time and guide the development of evidence-based patient management tools to improve outcomes.

## Key findings

### Question

To define the clinical status of patients infected by SARS-CoV-2 who require admission to an intensive care unit (ICU) and invasive interventions.

### Findings

The protocol of a pragmatic international, multicenter, observational clinical study of patients with confirmed or suspected SARS-CoV-2 infection admitted to ICUs around the world.

### Meaning

This is an evolving clinical registry, which will facilitate the characterization of patients and their management and provide real-time information on associate characteristics and outcomes. These data will assist clinicians in deriving evidence-based practices for the care of critically-ill patients infected by SARS-CoV-2.

## Clinical Trial Registration

ACTRN12620000421932. Available from: http://anzctr.org.au/ACTRN12620000421932.aspx.

## References

1 Zhu N, Zhang D, Wang W, et al. A Novel Coronavirus from Patients with Pneumonia in China, 2019. The New England journal of medicine. 2020;382(8):727–733.

2. The species Severe acute respiratory syndrome-related coronavirus: classifying 2019-nCoV and naming it SARS-CoV-2. Nature microbiology. 2020;5(4):536–544.

3. Dong E, Du H, Gardner L. An interactive web-based dashboard to track COVID-19 in real time. The Lancet Infectious diseases. 2020;20(5):533–534.

4. Barrasa H, Rello J, Tejada S, et al. SARS-CoV-2 in Spanish Intensive Care Units: Early experience with 15-day survival in Vitoria. Anaesthesia, criticalcare & pain medicine. 2020.

5. Grasselli G, Zangrillo A, Zanella A, et al. Baseline Characteristics and Outcomes of 1591 Patients Infected With SARS-CoV-2 Admitted to ICUs of the Lombardy Region, Italy. Jama. 2020;323(16):1574–1581.

6. Bhatraju PK, Ghassemieh BJ, Nichols M, et al. Covid-19 in Critically Ill Patients in the Seattle Region – Case Series. The New England journal of medicine. 2020.

7. Yang X, Yu Y, Xu J, et al. Clinical course and outcomes of critically ill patients with SARS-CoV-2 pneumonia in Wuhan, China: a single-centered, retrospective, observational study. The Lancet Respiratory medicine. 2020;8(5):475–481.

8. Wang D, Hu B, Hu C, et al. Clinical Characteristics of 138 Hospitalized Patients With 2019 Novel Coronavirus-Infected Pneumonia in Wuhan, China. Jama. 2020;323(11):1061–1069.

9. Ruan Q, Yang K, Wang W, Jiang L, Song J. Clinical predictors of mortality due to COVID-19 based on an analysis of data of 150 patients from Wuhan, Clinical predictors of mortality due to COVID-19 based on an analysis of data of 150 patients from Wuhan China. Intensive care medicine. 2020;46(5):846–848.

10. Vincent JL, Taccone FS. Understanding pathways to death in patients with COVID- 19. The Lancet Respiratory medicine. 2020;8(5):430–432.

11. Marini JJ, Gattinoni L. Management of COVID-19 Respiratory Distress. Jama. 2020.

12. Gattinoni L, Coppola S, Cressoni M, Busana M, Rossi S, Chiumello D. COVID-19 Does Not Lead to a “Typical” Acute Respiratory Distress Syndrome. American journal of respiratory and critical care medicine. 2020;201(10):1299–1300.

13. Klok FA, Kruip M, van der Meer NJM, et al. Incidence of thrombotic complications in critically ill ICU patients with COVID-19. Thrombosis research. 2020.

14. Levi M, Thachil J, Iba T, Levy JH. Coagulation abnormalities and thrombosis in patients with COVID-19. The Lancet Haematology. 2020.

15. Harris PA, Taylor R, Thielke R, Payne J, Gonzalez N, Conde JG. Research electronic data capture (REDCap)--a metadata-driven methodology and workflow process for providing translational research informatics support. Journal of biomedical informatics. 2009;42(2):377–381.

16. Wolkewitz M, Cooper BS, Bonten MJ, Barnett AG, Schumacher M. Interpreting and comparing risks in the presence of competing events. BMJ (Clinical research ed). 2014;349:g5060.

17. Steyerberg EW, Vickers AJ, Cook NR, et al. Assessing the performance of prediction models: a framework for traditional and novel measures. Epidemiology (Cambridge, Mass). 2010;21(1):128–138.

18. Lundberg SM, Erion G, Chen H, et al. From local explanations to global understanding with explainable AI for trees. Nature Machine Intelligence. 2020;2(1):56–67.

19. Moons KG, Altman DG, Reitsma JB, et al. Transparent Reporting of a multivariable prediction model for Individual Prognosis or Diagnosis (TRIPOD): explanation and elaboration. Annals of internal medicine. 2015;162(1):W1–73.

20. Bellani G, Laffey JG, Pham T, et al. Epidemiology, Patterns of Care, and Mortality for Patients With Acute Respiratory Distress Syndrome in Intensive Care Units in 50 Countries. Jama. 2016;315(8):788–800.

21. Rhodes A, Ferdinande P, Flaatten H, Guidet B, Metnitz PG, Moreno RP. The variability of critical care bed numbers in Europe. Intensive care medicine. 2012;38(10):1647–1653.

